# Teach, and teach and teach: does the average citizen use masks correctly during daily activities? Results from an observational study with more than 12,000 participants

**DOI:** 10.1101/2020.06.25.20139907

**Authors:** Evaldo Stanislau Affonso de Araújo, Fatima Maria Bernardes Henriques Amaral, Dongmin Park, Ana Paola Ceraldi Cameira, Murilo Augustinho Muniz da Cunha, Evelyn Gutierrez Karl, Sheila J. Henderson

## Abstract

COVID-19 is a new disease with no treatment and no vaccine so far. The pandemic is still growing in many areas. Among the core measures to prevent disease spread is the use of face masks. We observed 12,588 people in five Brazilian cities within the Baixada Santista metropolitan area. Even though this is densely populated region and heavily impacted by COVID-19 with a high risk population, only 45.1% of the observed population wore in face masks in a correct way, and another 15.5% simply did not use masks at all. The remainder used masks incorrectly, which is evidence of the worst scenario of people believing that they are protected when they are not. This is among the first studies, to the best of our knowledge, that measures real life compliance with face masks during this COVID-19 pandemic. It is our conclusion that it is paramount to first control the virus before allowing people back in the streets. We should not assume that people will wear masks properly. Equally important is to instruct and sensitize people on how to use face masks and why it is important.

## Introduction

The impact of COVID-19 has been enormous; everyone has had to change their social and health behaviors. Nonetheless, with every new day, COVID-19 becomes an easier and less frightening subject for the less informed. Especially with the need for daily income, the lack of social support for the stay at home policy, and in some countries like Brazil, the political misguidance contrary to the recommendations of the Science and Health Authorities [1], public pressure has mounted to end the quarantine. Some Brazilian cities are arranging the progressive reopening of commercial areas and allowing people to move freely about town. As the quarantine ends, with the numbers of cases and deaths on the rise, it has now become obligatory to wear a face mask.

The Baixada Santista metropolitan area is an immense harbor region (the largest in Latin America), providing an oil and industrial area close to São Paulo, with a large retired population of 60+ years in age. COVID-19 is highly prevalent with rising deaths (1,706 cases/100,000 inhabitants and 65.3 deaths/100,000, as of June 20, 2020, for the city of Santos) [2]. Although a population-based serologic survey conducted in the region every two weeks in the last two months (since June 11, 2020) showed a variation in anti-SARS-CoV-2 antibodies from 1.4 to 6.6%, the region still eased on the Social Distancing Rules. Due to the strategic nature of the region, its aging population, and the growing numbers of cases, and antibodies prevalence, the impact of COVID-19 is growing rapidly and perilously. For a proper risk assessment, it is critical to understand the degree of public compliance with protective measures. The face mask is a cornerstone measure to protect against the COVID-19 infection [3]. To evaluate if, how often, and in what way people wear their face masks, we conducted an observational study in five major cities in the Baixada Santista metropolitan area with a sample of over 12,000 observations.

## Material and Methods

From June 17th to 19th, 2020, medical student researchers went to large commercial streets in the cities of Santos, São Vicente, Cubatão, Guarujá, and Praia Grande. For three consecutive days, for a period of one hour, the same researcher occupied the same spot on the same street, at the same time, and observed and recorded if, how many, and in what way, people were wearing their face masks. The sample size was based on the numbers of people at streets seen and recorded during the specified hours of observation at the specified place, which reflected the real life scenario for an average weekday. To guard against observer bias, the researchers were included in determining the categories of face mask use, and we ensured consensus about these categories before the study began. The following categories were observed: people wearing masks covering mouth and nose, firmly adjusted; or individuals wearing a mask with their nose and/or mouth exposed; people not wearing masks; others were wearing a poorly fitting mask; and, finally, people who touch their masks during use. Results were plotted and analyzed in total and by city.

## Results

Table 1 (by number and percentage) and Figure 1 (by percentage) display the research results by city and in total. Overall, an average of 45.1% of the people observed wore their masks properly, within a city by city range of 39.1% to 63.5%. Within the remaining 54.9%, 15.5% wore no mask (ranging from 12.7% to 18.8%), 12.9% wore masks but exposed their mouth and nose (range: 9.9% to 17.6%), 12.0% exposed their nose only (7.9% to 16.6%), 17.8% touched their mask during use (0.0% to 14.0%), and 6.5% wore poorly fitted masks (1.2% to 10.7%). The number of observations across the five cities was similar from 2,270 people (18.0%) in São Vicente to 3055 (24.6%) in Santos.

**Table 1:** Observations of face mask compliance by city and region

| City | Date in June 2020 | Correct Use | No mask | Exposed Mouth and Nose | Exposed Nose | Touching Face During Use | Poorly Fitted Mask | City Total | % |
| --- | --- | --- | --- | --- | --- | --- | --- | --- | --- |
| CUBATAO | 17th | 640 | 130 | 104 | 62 | 0 | 14 |  |  |
|  | 18th | 604 | 120 | 68 | 50 | 0 | 10 |  |  |
|  | 19th | 266 | 152 | 76 | 76 | 0 | 5 |  |  |
| **TOTAL CUBATAO** |  | **1,510** | **402** | **248** | **188** | **0** | **29** | **2,377** | **18.8%** |
| % |  | 63.5% | 16.9% | 10.4% | 7.9% | 0% | 1.2% | 100% |  |
| PRAIA GRANDE | 17th | 360 | 108 | 76 | 66 | 38 | 46 |  |  |
|  | 18th | 318 | 170 | 194 | 108 | 40 | 92 |  |  |
|  | 19th | 411 | 188 | 96 | 62 | 53 | 42 |  |  |
| **TOTAL PRAIA GRANDE** |  | **1,089** | **466** | **366** | **236** | **131** | **180** | **2,468** | **19.6%** |
| % |  | 44.1% | 18.8% | 14.8% | 9.5% | 5.3% | 7.3% | 100% |  |
| SANTOS | 17th | 321 | 145 | 66 | 174 | 179 | 146 |  |  |
|  | 18th | 351 | 130 | 161 | 103 | 116 | 73 |  |  |
|  | 19th | 349 | 172 | 141 | 228 | 135 | 65 |  |  |
| **TOTAL SANTOS** |  | **1,021** | **447** | **368** | **505** | **430** | **284** | **3,055** | **24.6%** |
| % |  | 33.4% | 14.6% | 12.0% | 16.6% | 14.0% | 9.3% | 100% |  |
| GUARUJA | 17th | 198 | 107 | 138 | 114 | 80 | 101 |  |  |
|  | 18th | 295 | 71 | 161 | 103 | 65 | 61 |  |  |
|  | 19th | 304 | 170 | 128 | 126 | 98 | 98 |  |  |
| **TOTAL GUARUJA** |  | **797** | **348** | **427** | **343** | **243** | **260** | **2,418** | **19.2%** |
| % |  | 32.9% | 14.3% | 17.6% | 14.1% | 10.0% | 10.7% | 100% |  |
| SAO VICENTE | 17 | 447 | 97 | 79 | 79 | 58 | 27 |  |  |
|  | 18 | 384 | 96 | 78 | 78 | 50 | 23 |  |  |
|  | 19 | 436 | 96 | 68 | 82 | 71 | 21 |  |  |
| **TOTAL SAO VICENTE** |  | **1,267** | **289** | **225** | **239** | **179** | **71** | **2,270** | **18.0%** |
| % |  | 55.8% | 12.7% | 9.9% | 10.5% | 7.8% | 3.1% | 100% |  |
| **Region TOTAL** |  | **5,684** | **1,952** | **1,634** | **1,511** | **983** | **824** | **12,588** | **100%** |
| % |  | **45.1%** | **15.5%** | **12.9%** | **12.0%** | **7.8%** | **6.5%** | **100%** |  |

**Figure 1.**
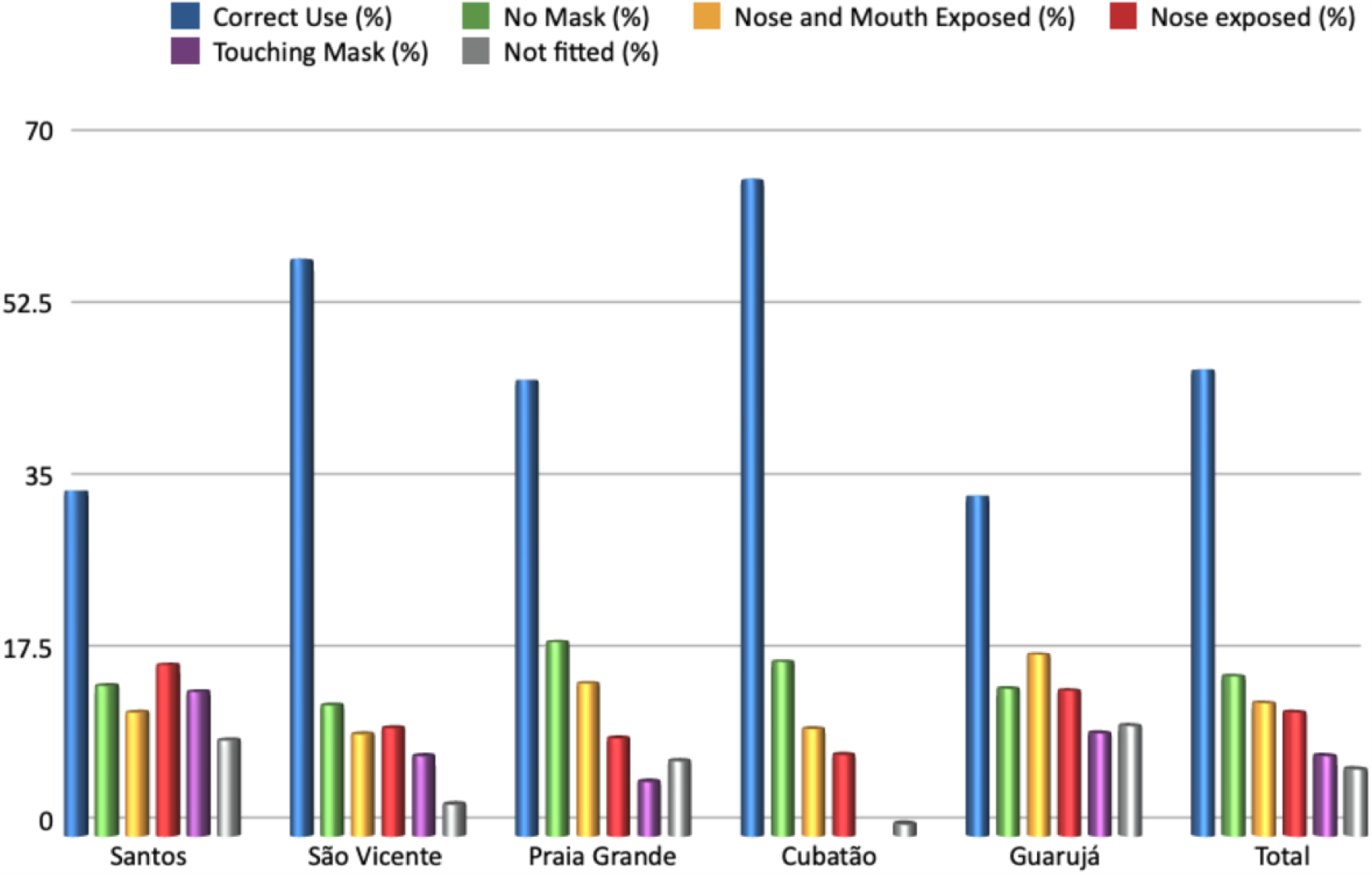
Relative use of face masks by city and region (%)

## Discussion

At the present time, there is little, if any, research data on face mask compliance worn to prevent the spread of COVID-19. In this study, over three consecutive days, we observed 12,588 people in the streets wearing, or not, masks in their routine lives. Overall, only 45.1% of the people observed wore their masks properly, 39.4% wore their masks improperly, and 15.5% did not wear masks at all. The 39.4% who were wearing masks improperly were likely to be as dangerous as those not wearing a mask. This suggests that many mask-wearing people were feeling safe, but in reality, were not, and therefore risking presence in the streets in an inadvisable fashion.

Non-pharmacological measures are the main line of defense against COVID-19 [3]. Several barriers—physical or behavioral—should exist between the susceptible host and SARS-CoV-2. Face masks have grown in relevance during this pandemic, particularly going from health care facilities to the average people daily activities. Face masks are now a cornerstone in prevention, no matter who or where the need exists. Nevertheless, face masks are new for the public. Face masks are not comfortable to wear, and they require a new routine for how to wear, how to remove, how to preserve and clean, and so forth. Many people complain of a lack of oxygen while wearing masks. Therefore, it may be overly optimistic to count on public compliance with face masks as a tool to prevent infection and as a measure to make it safe to relax hard social distancing.

Our results are quite disturbing. We found that only 45.1% of people wore face masks properly and safely in a research sample observed in a region with significant prevalence of COVID-19, among people at high risk of infection (because of poverty and high prevalence) and complications (due to age and pre-existing health conditions). According some mathematical models, more than 80% of people must wear a face mask properly for efficacy in COVID-19 spread prevention [4]. As a public health measure and the successful defense against the spread of COVID-19, the use of masks to protect the public, in lieu of quarantine, depends on each individual adhering to proper use [5]. We must ask ourselves: *who are we deceiving with the false belief of high compliance?* Before we move to some plan of further relaxing control measures, it is clear that safe social distancing is still a critical target to be achieved in order to reduce circulation of the virus. These research results from Baixada Santista, revealing inadequate public compliance, suggest that we are playing with fire and jeopardizing people lives, and quite literally, offering them over to the virus. Prevention of COVID-19, like some other infectious diseases, (so far) lacks an effective vaccine and therefore requires a change in public behavior. HIV taught us how hard is to avoid risk with a single behavioral change, such as using a condom. So, before we move ahead with peace of mind, and erroneously assume that people will wear masks routinely and properly, it is paramount that we teach, and teach, and teach use of face masks, and explain why one’s life depends on proper use.

To the best of our knowledge, this study is the first to observe COVID-19 prevention from the perspective of actual observed compliance. This was an observational study, in a single region with no intervention. The generalizability of this study is unknown, however, this study can be easily replicated at low cost in other regions around the world. Observational studies are useful as preliminary but rapid research to raise significant red flags in current efforts against COVID-19.

## Data Availability

The data that support the findings of this study are available from the corresponding author, Evaldo Stanislau Affonso de Araujo, upon reasonable request.

## Author Contributions

Evaldo Stanislau Affonso de Araújo [a]

Fatima Maria Bernardes Henriques Amaral [b]

Dongmin Park [b]

Ana Paola Ceraldi Cameira [b]

Murilo Augustinho Muniz da Cunha [b]

Evelyn Gutierrez Karl [b]

Sheila J. Henderson [c]

[a] concept of the study, wrote manuscript, prepared dataset

[b] field observation

[c] reviewed and prepared manuscript

## References

1. Editorial. COVID-19 in Brazil: “So what?”. Lancet, 2020, 339: 1461, May 9, 2020. https://doi.org/10.1016/S0140-6736(20)31095-3

2. ESPECIAL COVID-19 – Dados por Município, 2020. https://www.brasil.io/covid19/ Accessed 20th June 2020

3. DK Chu, EA Akl, S Duda, K Solo, S Yaacoub, et al. Physical distancing, face masks, and eye protection to prevent person-to-person transmission of SARS-CoV-2 and COVID-19: a systematic review and meta-analysis. Lancet, In press. https://doi.org/10.1016/S0140-6736(20)31142-9.

4. SE Eikenberry, M Mancuso, E Iboi, T Phan, K Eikenberry, et al. (2020). To mask or not to mask: Modeling the potential for face mask use by the general public to curtail the COVID-19 pandemic. Infectious Disease Modelling, 5: 293-306, 2020. https://doi.org/10.1016/j.idm.2020.04.001

5. WHO. Advice on the use of masks in the context of COVID-19. Interim guidance. 5 June 2020. https://www.who.int/publications/i/item/advice-on-the-use-of-masks-in-the-community-during-home-care-and-in-healthcare-settings-in-the-context-of-the-novel-coronavirus-(2019-ncov)-outbreak Accessed 20th June 2020

